# Predicting the efficacy of Recombinant Human Thrombopoietin in Treating Cancer Therapy-Related Thrombocytopenia:based on stacking ensemble methods

**DOI:** 10.64898/2025.12.14.25342230

**Authors:** Kun Hou, Li Feng, Haiwen Lu, Zhenfei Wang

## Abstract

The project aimed to develop a data-driven approach for predicting platelet recovery in cancer treatment–induced thrombocytopenia (CTIT) patients receiving recombinant human thrombopoietin (Rh-TPO). By integrating key clinical indicators into a predictive modeling framework, the study sought to enhance understanding of individual treatment responses and facilitate timely clinical decision-making. A retrospective two-stage modeling analysis was conducted on 400 hospitalized CTIT patients who received Rh-TPO therapy in 2023, with data randomly split into training and testing sets. Following rigorous feature selection, multiple machine learning regression models were trained, and a stacking ensemble model was developed to leverage their combined strengths. Model performance was evaluated on the test set, and an explainable AI analysis was subsequently performed to identify the most influential clinical features driving platelet recovery. Eight variables—baseline platelet count before Rh-TPO initiation, duration of Rh-TPO therapy, pre-anticancer treatment platelet count, timing of follow-up blood tests, hemoglobin level, age, height, and ethnicity—were consistently identified as key predictors. The stacking ensemble model achieved the highest predictive accuracy, demonstrating superior agreement between predicted and observed platelet recovery trajectories compared to individual models. Improved prediction stability was associated with more comprehensive pretreatment assessments and well-defined treatment timelines. These results underscore the value of integrating heterogeneous clinical data through ensemble learning and suggest that such predictive models could support earlier identification of suboptimal platelet recovery, thereby improving CTIT management.

## 1. Introduction

Cancer Treatment-Induced Thrombocytopenia (CTIT) is a common adverse reaction associated with anti-cancer drugs, affecting up to 15% to 25% of patients undergoing anti-tumor therapy [1,2], with grade III/IV cases occurring at a rate of approximately 3% [3]. CTIT not only increases the risk of bleeding, prolongs hospital stays, and raises medical costs, but it can also negatively impact anti-tumor therapeutic efficacy and potentially lead to patient mortality [4].To reduce or prevent the survival risks posed by CTIT, various domestic and international guidelines and consensus statements, in conjunction with the evolving landscape of anti-tumor and platelet-boosting therapeutic agents, have refined the standard prevention and treatment strategies for CTIT and established corresponding standards [5,6,7]. For example, the Chinese Society of Clinical Oncology (CSCO) guidelines recommend the use of Recombinant Human Thrombopoietin Injection (Rh-TPO) to prevent and treat CTIT, aiming to reduce the decline in platelet counts following anti-cancer drug treatment, shorten the duration of thrombocytopenia, and decrease the frequency of platelet transfusions [8,9]. As Rh-TPO is categorized as a Class B reimbursable drug under the medical insurance system, it has become the primary choice for the clinical management of CTIT in China. However, despite these guidelines and recommendations, several challenges and uncertainties remain in clinical practice, including individual patient variability, regional differences, cancer types, and treatment regimens, all of which can significantly influence the effectiveness of Rh-TPO in treating CTIT.

Therefore, developing an effective predictive model to forecast the platelet counts improvement in patients with CTIT before initiating Rh-TPO treatment is crucial for achieving personalized treatment and optimizing clinical decision-making. Traditional data analysis methods, however, face significant limitations in this domain, such as their inability to efficiently handle multi-dimensional complex data, the presence of substantial human bias, and the lack of dynamic updating and real-time prediction capabilities.

Machine learning methods are a promising data statistical analysis technique in the field of medicine and an essential component of artificial intelligence. Compared with traditional statistical methods, machine learning can mine large datasets for fitting, enhance data processing capabilities, uncover data patterns, and improve the precision and efficiency of multi-dimensional data processing [10,11].In recent years, the use of machine learning algorithms to establish disease treatment-related predictive models has garnered increasing attention [12]. Machine learning has made significant advancements in predicting the occurrence risk of tumor treatment-related complications, assessing the effectiveness of anti-tumor therapy, and mining and validating tumor molecular markers [13,14,15]. However, no corresponding research has been conducted on predicting the efficacy of Rh-TPO treatment for CTIT. Therefore, this study aimed to develop a predictive model for the efficacy of Rh-TPO treatment for CTIT using clinical data from patients treated at our hospital and machine learning and stacking algorithms .This approach is significant for improving the therapeutic effects of Rh-TPO on CTIT, reducing hospitalizations and emergency interventions due to thrombocytopenia, thereby increasing the efficiency of medical resource utilization and enhancing both the safety and effectiveness of anti-tumor treatments for cancer patients.

## 2. Methods

### 2.1 Data sources and study population

This retrospective study included the cases of patients who developed CTIT and received Rh-TPO treatment from 01/01 2023 to 31/12/2023.Patients under the age of 18, incomplete case records, and those without dynamic platelet counts monitoring were excluded. Data on patient demographics, cancer treatment details, and platelet counts trends were extracted to create the dataset. Due to the retrospective nature of the study, informed consent was waived. The study was approved by the Ethics Committee of Inner Mongolia Medical University.The study primarily collected variables,including the platelet counts of patients prior to cancer therapy, days of post cancer therapy follow-up, and the platelet counts before Rh-TPO administration. Additionally, data were gathered regarding the dosage and duration of Rh-TPO administration, as well as the platelet counts recorded after completing Rh-TPO treatment, in conjunction with other relevant parameters included baseline alanine aminotransferase (ALT) , baseline aspartate aminotransferase (AST), baseline serum creatinine, baseline hemoglobin concentration, height, weight, gender, ethnicity, and age.The study was approved by the Ethics Committee of Inner Mongolia Medical University with the approval numberYKD202401033.The study was approved on March 2024.Informed consent was not required because all protected health in the database was de-identified and did not influence clinical care. The requirement for informed consent was waived by the Medical Ethics Committee of Inner Mongolia Medical University with the approval numberYKD202401033.All methods in our study were performed strictly in accordance with the Declaration of Helsinki.

The primary outcome measure was the platelet counts after completing Rh-TPO treatment.The rationale for this approach was based on several variables: platelet counts before cancer therapy, days of post cancer therapy follow-up, platelet counts before Rh-TPO administration, dosage and duration of Rh-TPO administration, baseline alanine aminotransferase (ALT),baseline aspartate aminotransferase (AST),baseline serum creatinine,baseline hemoglobin,height, weight, gender, ethnicity, and age.

### 2.2 Data Preprocessing and Features Selection

Variables with a missing data rate exceeding 30% were excluded from the analysis. For the remaining variables,missing values were imputed using the K-Nearest Neighbors (KNN) algorithm. All variables indices underwent preprocessing to ensure that non-integer data were converted to integer format. Categorical variables such as gender and ethnicity were encoded as follows: Han = 0, Mongolian = 1, Bai = 2, Manchu = 3; Female = 1, Male = 0. After preprocessing, the original dataset was organized for subsequent analysis and modeling. The dataset was then randomly partitioned into training and test sets in a ratio of 7:3.

In this study, we employed the least absolute shrinkage and selection operator (LASSO)model regression to identify the most significant features for our predictive model. This approach not only reduces model complexity but also mitigates the risk of overfitting. We utilized the LassoCV model to perform 3-fold cross-validation on the training set across various values of the regularization parameter alpha. The optimal alpha values were determined by analyzing mean squared error (MSE) plots and coefficient path plots. Based on these optimal values, we retained features with non-zero coefficients as important predictors while eliminating those with zero coefficients. Ultimately, the selected features enhance both the predictive performance and interpretability of the model while simplifying its structure.

### 2.3 Independent ML Model Development

We constructed six independent machine learning models: Support Vector Machine Regressor (SVM), Random Forest Regressor (RF), Light Gradient Boosting Machine (LightGBM), Extreme Gradient Boosting Regressor (XGBoost), Categorical Boosting Regressor (CatBoost), and Categorical Boosting with Genetic Algorithm Regressor (CatBoost-GA).Each model was trained on the training set, and predictions and performance evaluations were conducted on the test set. Given that platelet count is a continuous variable, model performance was assessed using the following metrics: coefficient of determination (R² score), mean squared error (MSE), mean absolute error (MAE), root mean square error (RMSE), and mean absolute percentage error (MAPE).

### 2.4 Stacking Ensemble Learning

To enhance the accuracy and generalization ability of the final prediction model, this study employed a stacking algorithm to integrate multiple base models, thereby improving prediction accuracy. The performance of this algorithm has been demonstrated to surpass that of boosting and bagging ensemble regression algorithms. Specifically, we selected the three best-performing models from the six independent models constructed in the previous step as base models, each configured with finely tuned hyperparameters. Subsequently, the predictions from these base models were used as input features for a linear regression model (Linear Regression) as the final stacking ensemble model. The training of this stacking ensemble model was conducted using 3-fold cross-validation to enhance its generalization ability and prevent overfitting. Finally, we compared the performance of the six independent models with that of the final stacking ensemble model.y. The modeling process of this study was shown in Fig 1.

**Figure 1.**
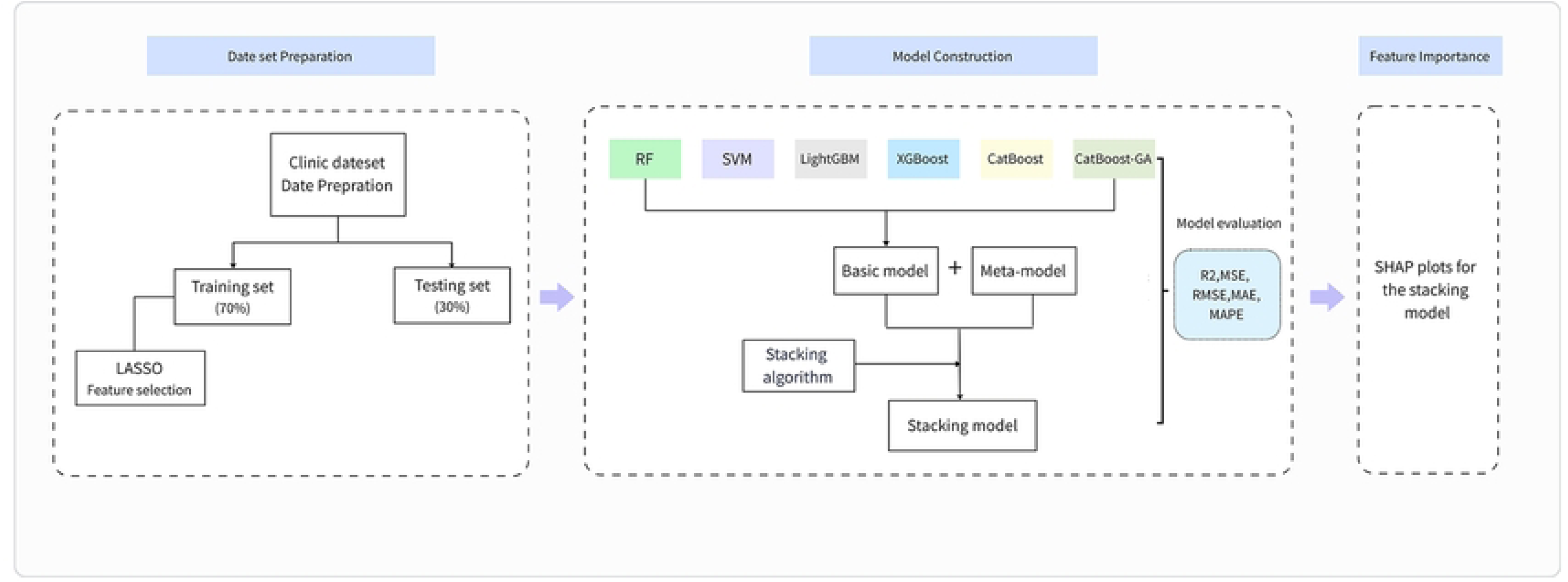
The modeling process of this study. LASSO, the least

### 2.5 Model interpretation

This study employs the SHapley Additive exPlanations (SHAP)method to interpret model results, presenting both SHAP summary plots and average SHAP values plots. In the SHAP summary plot, the Y-axis represents features, while the X-axis indicates the impact of features on outcomes. Each point represents a sample, with red indicating high-risk values and blue indicating low-risk values. The modeling process of this study was shown in Fig 1.

### 2.6 Statistical analysis

Machine learning model training and testing were conducted in the Python 3.7.4 environment. Statistical analysis of clinical data was performed using SPSS 27.0. Continuous variables were expressed as mean ± standard deviation (SD) when the data were normally distributed; for non-normally distributed data, continuous variables were expressed as median with first and third quartiles [M(P25, P75)]. Categorical data were presented as the number of cases and percentages (%).

## 3. Results

### 3.1 Baseline characteristics

A total of 400 cases had been included in the study, with patient datas covering age, gender, ethnicity, height, weight, platelet counts before cancer therapy (PLTs before cancer therapy),days of post cancer therapy follow-up,platelet counts before Rh-TPO administration(PLTs before Rh-TPO), platelet counts after Rh-TPO administration(PLTs after Rh-TPO), duration of Rh-TPO,as well as the the baseline alanine aminotransferase(ALT), aspartate aminotransferase (AST), serum creatinine, hemoglobin levels. These were all used as features for model building. As shown in Table 1,the study found that among the 400 tumor patients with CTIT, the majority were elderly women over 60 years old, and Han patients were in the majority, but patients from other ethnic minorities also accounted for a certain proportion. Before anti-tumor treatment, the platelet count of all patients was within the normal range; before using Rh-TPO, the median platelet count of the patients was 59.8×10^9^/L. The median treatment course of Rh-TPO was 5 days.

### 3.2 Lasso regression for feature variable selection

The dataset contained no missing data, eliminating the need for imputation. The results of LASSO regression on feature screening were shown in Fig 2.We identifyed seven feature variables with non-zero coefficients from the training set, specifically: ethnicity, height (cm), baseline creatinine, platelet counts before cancer therapy,days of post cancer therapy follow-up,platelet counts before Rh-TPO administration (PLTs before Rh-TPO), and duration of Rh-TPO treatment. These feature variables were utilized as inputs for subsequent model training.

**Figure 2.**
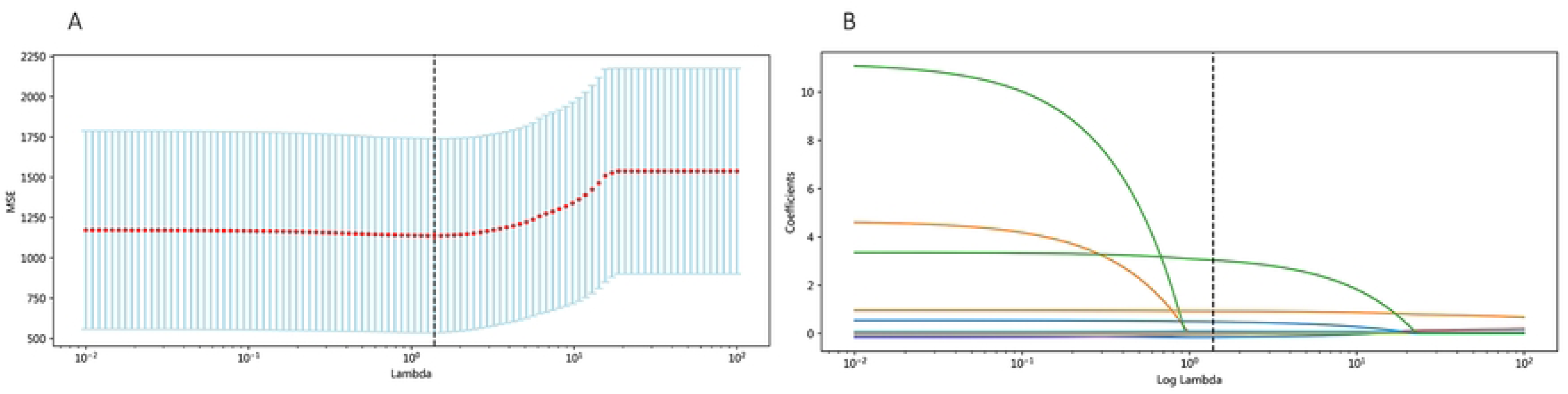
Feature selection using the least absolute shrinkage and

### 3.3 Model evaluation and performance

RF,XGB,and CatBoost exhibited the best performance. Therefore, we selected these three models as base models and used Linear Regression as the meta-model for the stacking learning.The prediction performance of the six independent models and the stacking ensemble model is presented in Table 2. The predictive ability of the stacking ensemble model on the training and testing set is illustrated in Fig 3.A comparison of the comprehensive performance of each model is shown in Fig 4. The performance of the stacking model was superior, as evidenced by the highest R² and the lowest MAE, MSE, RMSE, and MAPE.

**Figure 3.**
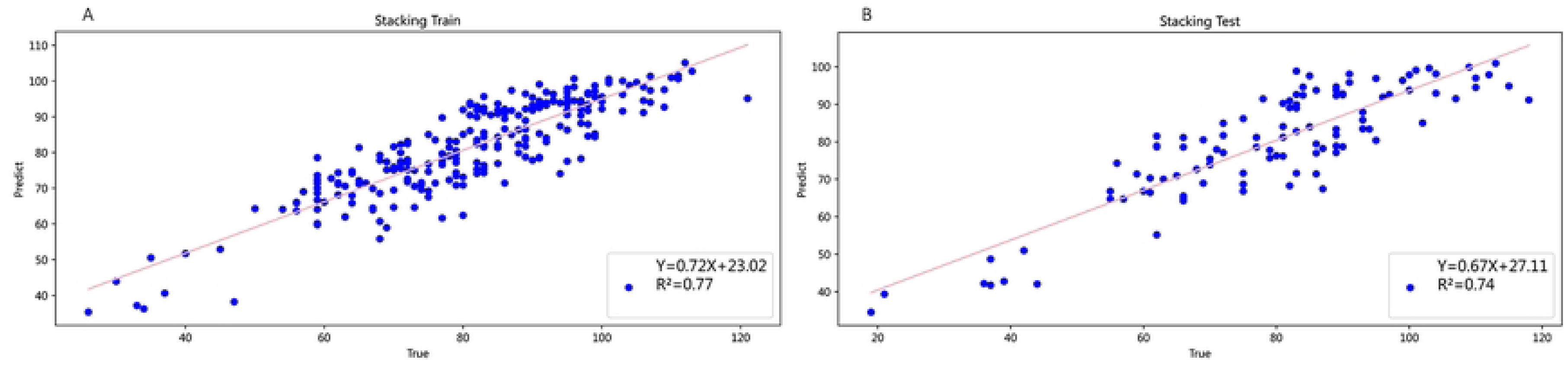
The predictive ability of the stacking ensemble model

**Figure 4.**
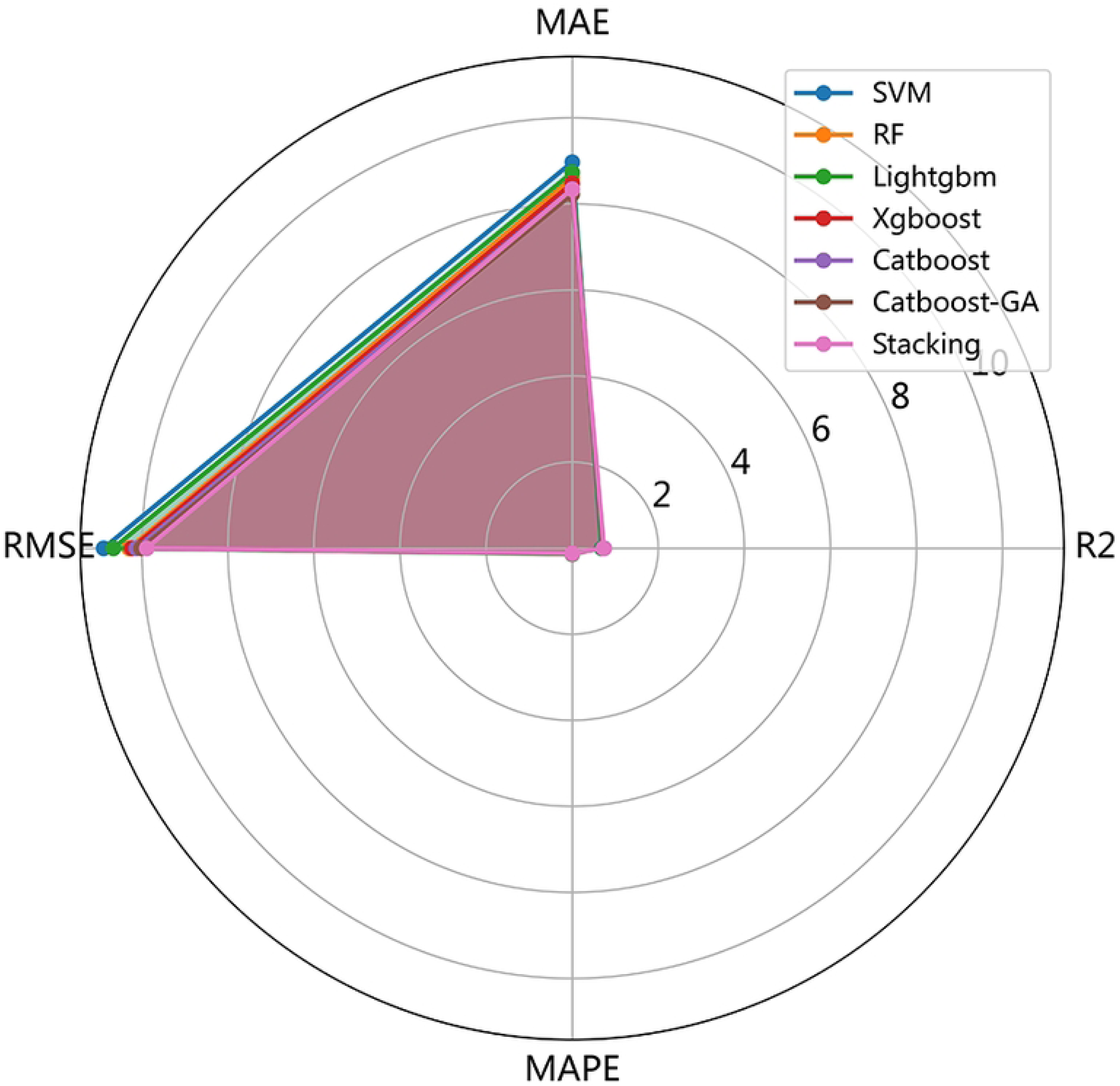
The radar chart for comprehensive comparison of 7 m

### 3.4 Model interpretation

After model evaluation, the stacking ensemble model was finally selected as the optimal model. The feature importance of the optimal model was ranked based on SHAP values. Fig 5A shows the average SHAP values of the seven features, while Fig 5B illustrates their importance ranking. From Fig 5A, the overall impact of each feature on the model output can be intuitively observed. The larger the average SHAP value, the more significant the feature’s impact on the prediction result. In Fig. 5B, the position of each feature on the Y-axis from top to bottom indicates a decreasing relative importance to the model prediction results, with the most important feature at the top. The X-axis represents the SHAP value, which measures the contribution of each feature to the model’s predictions. Positive SHAP values indicate that the feature enhances the predicted result (i.e., increases the predicted value), while negative SHAP values indicate that the feature inhibits the predicted result (i.e., decreases the predicted value). The color of the points represents the original value of the feature, with red indicating high values and blue indicating low values. If a feature has a positive SHAP value and is colored red, it means that the feature increases the prediction result at high values. Conversely, if it has a negative SHAP value and is colored blue, it means that the feature reduces the prediction result at low values.Therefore, the importance rankings of the seven characteristic variables are as follows: PLTs before Rh-TPO, duration of Rh-TPO treatment, baseline creatinine, PLTs before cancer therapy, days of post-cancer therapy follow-up, height, and ethnicity. Notably, higher PLTs before Rh-TPO administration and longer durations of Rh-TPO treatment are associated with better platelet improvement effects after Rh-TPO treatment for CTIT.

**Figure5A.**
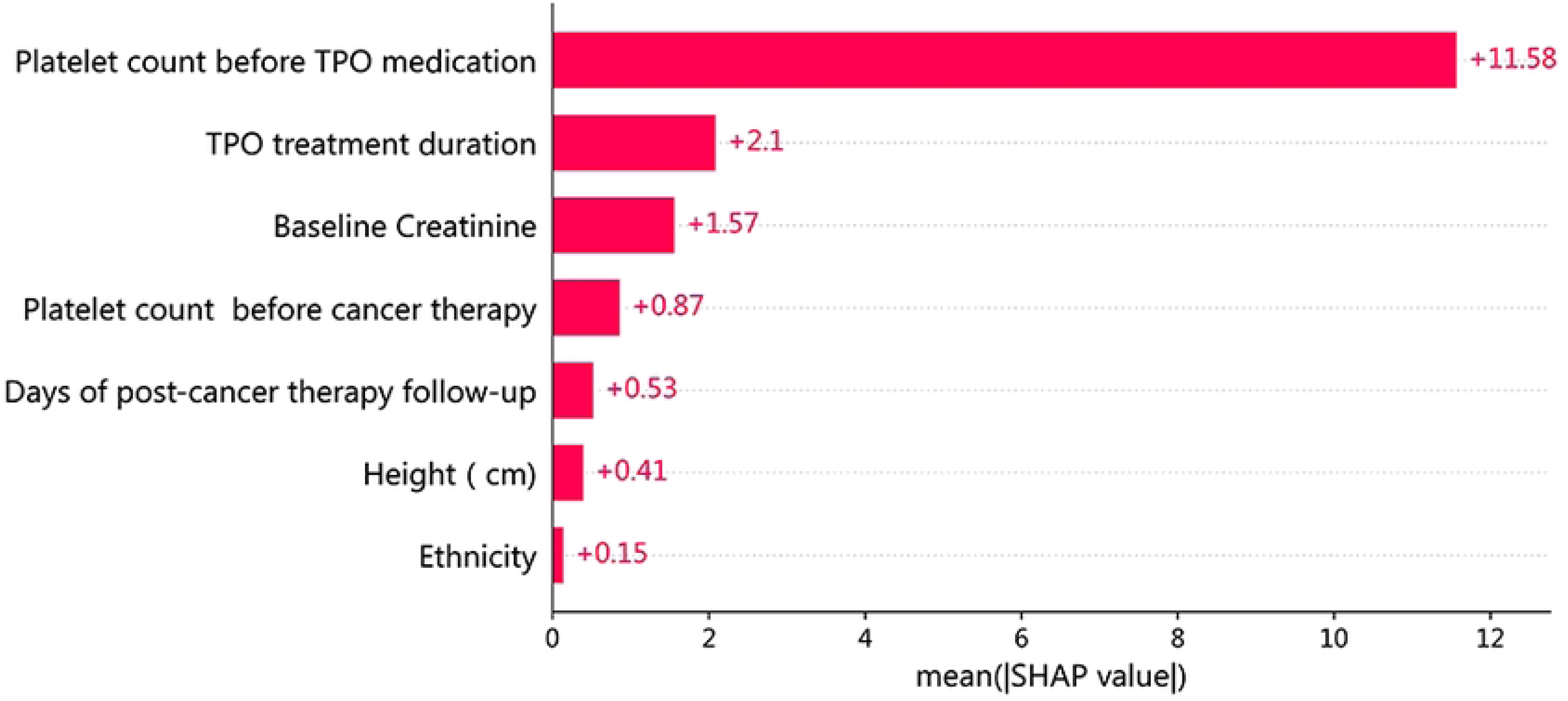
The ranking of the average SHAP values of the 7 featur

**Figure5B.**
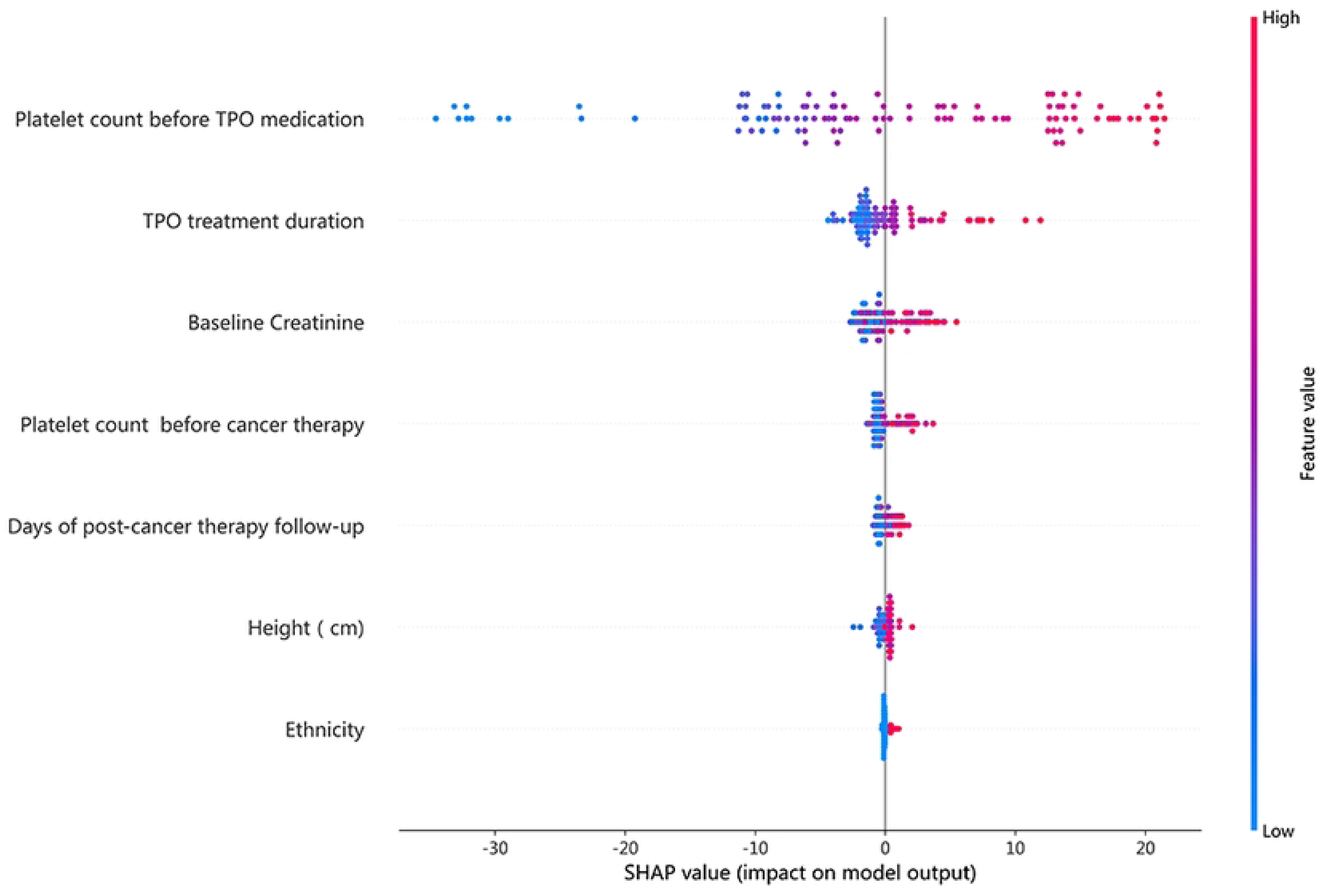
The ranking of the importance of 7 features in the stac

### 3.5 Visual interface

To achieve real-time prediction of Rh-TPO treatment effects, we developed an interactive web application using Streamlit.The website address is https://platelet-qigobpmrnyevbrnyfkcftm.streamlit.app/. Users can input specific patient characteristics on the web page, such as age, hemoglobin level, and other relevant parameters, and the system will predict the patient’s platelet value after a specified period based on the final prediction model. The application features a simple and intuitive interface, enabling users to easily perform predictions and obtain results quickly. This provides a user-friendly platform for subsequent external validation of our study.

## 4. Discussion

In recent years, remarkable progress has been made in the application of machine learning models in the prediction of the efficacy of antineoplastic drugs. Among the many prediction models, Ensemble Learning method is particularly attractive, especially the Stacking method, because it can effectively combine the advantages of different models to improve the prediction performance, and is widely used in a variety of cancer therapy-related tasks.In this study, the research subjects is the platelet counts following Rh-TPO treatment, which is a continuous variable. The objective of the predictive model was to minimize the discrepancy between predicted and actual measured values, thereby classifying its performance as a regression problem. Consequently, we selected R², MSE, MAEas the evaluation metrics for model performance. It is widely acknowledged in the literature that a model demonstrates optimal predictive performance when the R2 value for the training set ranges from 0.8 to 1 and the R2 value for the validation set lies between 0.5 and 1, while minimizing MSE and MAE further enhances model accuracy. Despite the relatively small test set of only 120 cases, the stacking ensemble model achieved R² values exceeding 0.7 on the test set, demonstrating robust fitting performance. Compared to six independent models, the stacking ensemble model exhibited the highest R² value and the lowest error. This indicates that the application of machine learning, particularly stacking methods, not only enhances the accuracy of tumor drug response prediction but also highlights the advantages of stacking in addressing complex problems. Unlike traditional single models, the stacking method improves overall prediction performance by weighting and adjusting the outputs of multiple base models. Based on our experimental results, the stacking method significantly enhanced the accuracy of drug response prediction while reducing the model’s sensitivity to individual features or data noise [16]. When selecting base models, XGBoost and CatBoost emerge as two of the most prominent gradient boosting tree (GBDT) algorithms in contemporary machine learning. XGBoost excels in handling non-linear data, capturing feature interactions, processing missing values, and implementing model regularization [17]. In contrast, CatBoost demonstrates exceptional proficiency in managing categorical variables through its innovative sorting method and efficient encoding techniques, enabling it to perform remarkably well across a variety of complex real-world problems [18]. By leveraging these two models as base learners, we can fully capitalize on their respective strengths for diverse tasks. Despite its relative simplicity, linear regression is employed as a meta-learner in stacking methods. Its advantages include reducing overfitting and enhancing generalization capabilities, particularly when aggregating predictions from multiple base models; these benefits are even more pronounced in such contexts. In this research, we discovered that employing linear regression as a meta-learner to combine the outputs of multiple base models successfully mitigated the bias and variance issues of each model, leading to notable improvements in both prediction stability and accuracy. The inherent simplicity and interpretability of linear regression provided us with valuable insights into how different base models contribute to the final prediction outcomes. When compared to more complex deep learning models or other highly nonlinear meta-learners, the performance of linear regression in this context was surprisingly effective.We propose that the role of linear regression within stacking ensemble models goes beyond simply acting as a weighting mechanism. Instead, it fosters collaborative interactions among the predictions from various base models by leveraging learned relationships, which helps to reduce potential errors associated with individual models.This approach allows for a more robust and accurate predictive framework, enhancing the overall reliability of the ensemble method [19,20,21].

In clinical settings, the balance between predictive performance and interpretability must be carefully considered when applying models. Therefore, interpreting feature contributions is particularly important. Based on SHAP,we conducted multiple analyses and final ranking of feature importance. Among these features, the platelet counts before Rh-TPO treatment ranked highest, with its average SHAP value significantly higher than the other six features, indicating its greatest contribution to the prediction model. This finding indirectly suggests that patients with higher pre-treatment platelet counts may experience better responses to Rh-TPO in improving platelet counts associated with CTIT.Several published clinical studies support this idea, showing that patients with higher initial platelet counts recover more quickly after Rh-TPO treatment and have a lower risk of severe bleeding events [22,23,24,25]. Rh-TPO accelerates platelet production by promoting the proliferation and maturation of megakaryocytes, a process based on the hematopoietic potential of the patient’s own bone marrow. Low platelet counts prior to Rh-TPO treatment typically indicate a higher risk of bleeding, which can prolong the treatment duration and slow down platelet recovery. Additionally, factors such as a history of pelvic radiotherapy or chronic liver disease can further impact the efficacy and duration of Rh-TPO treatment. Therefore, understanding a patient’s pre-treatment platelet counts can assist physicians in making personalized treatment adjustments when utilizing Rh-TPO.During anti-tumor drug therapy, clinicians and clinical pharmacists should conduct a comprehensive evaluation of CTIT risk, considering the patient’s medical history and previous treatments, while regularly monitoring platelet counts. When platelet counts fall to the lower limit of normal values, a thorough assessment is warranted; patients identified as at risk for bleeding should promptly initiate Rh-TPO therapy.The second most significant characteristic associated with SHAP values is the duration of Rh-TPO treatment. Prolonged administration of Rh-TPO has been correlated with more favorable improvements in patients’ platelet counts. Rh-TPO promotes the final production and release of platelets by stimulating megakaryocytes in the bone marrow, a process that typically takes several days to weeks and varies among individuals. Consequently, the therapeutic effect of Rh-TPO is not immediate, and the duration of treatment directly influences the timeline for observing therapeutic benefits. If the treatment cycle is brief, patients may not fully capitalize on the effects of Rh-TPO, resulting in only modest or transient improvements in platelet levels. A longer treatment duration often allows more time for the drug to exert its effects, thereby facilitating ongoing platelet production and enabling patient platelet counts to gradually return to within normal ranges. Especially for patients with thrombocytopenia caused by chemotherapy, bone marrow suppression, or other underlying conditions, the rate of platelet consumption is significantly increased. Short-term Rh-TPO treatment may only address immediate platelet deficiency needs, whereas a prolonged course can help maintain stable platelet levels and reduce complications associated with thrombocytopenia, such as bleeding. In cases of chronic or recurrent thrombocytopenia, the duration of Rh-TPO therapy directly influences the speed and extent of platelet recovery. Therefore, extending the treatment period may be particularly beneficial for patients with persistent or recurrent conditions. However, it is important to note that long-term use of Rh-TPO may increase the risk of thrombosis and induce immune system changes [26]. Consequently, the treatment regimen should be individualized based on the patient’s specific clinical condition. While a longer treatment course can enhance therapeutic efficacy, careful consideration must be given to balancing risks and benefits.

## 5. Limitations

This study has several limitations. Firstly, the dataset used is retrospective in nature, which may limit the model’s generalization ability. Future research should aim to enhance the model’s generalizability by incorporating more diverse and prospective data sources. Secondly, the amount of data included in this study is relatively limited. Expanding the dataset is crucial for increasing the frequency of machine learning iterations and enhancing data processing capabilities, thereby further optimizing the model’s accuracy. In future work, we plan to explore integrating more complex meta-learners (such as deep learning models) with traditional linear regression. By leveraging advanced feature engineering and hyperparameter optimization techniques, we aim to further improve the performance of the stacking method. Through these efforts, we expect to provide more accurate tools for tumor drug response prediction and personalized treatment.

## 6. Conclusion

The findings of this study indicate that the stacking ensemble model demonstrates commendable performance in predicting the efficacy of Rh-TPO treatment for chemotherapy-induced thrombocytopenia (CTIT) in tumor patients. This provides valuable support for clinical decision-making. However, it is crucial to further collect relevant data to train and optimize the model, thereby enhancing the accuracy of predictive outcomes.

## Data Availability

All relevant data are within the manuscript and its Supporting Information files.

## Acknowledgements

Not applicable.

